# A Qualitative Descriptive Study Exploring Caregivers’ Information Needs and Experience Caring for a Child with Chronic Heart Failure

**DOI:** 10.1101/2024.02.27.24303476

**Authors:** Chentel Cunningham, Jennifer Conway, Ziad Zahoui, Mark Haykowsky, Shannon D. Scott

## Abstract

**Background:** Chronic phenotypes of pediatric heart failure pose life-long burdensome symptoms for the healthcare system and families. Treatment involves complex medical therapies with few surgical options until more advanced, refractory stages. Caregivers must become proficient in providing care to these vulnerable children in the home environment, which imposes a high amount of stress. Despite caregiver demands, little is known about caregiver information needs and experiences caring for a child with chronic heart failure. Therefore, a qualitative approach employing semi-structured interviews aimed to fill this knowledge gap.

**Methods and Results:** A qualitative descriptive methodology guided our study. Participants were recruited from a tertiary cardiac centre in Edmonton, Alberta, Canada. Data collection and analysis occurred concurrently. Semi-structured interviews were conducted until data redundancy was achieved. Inductive content analysis was used to uncover categories. Eleven interviews identified five main categories. Three categories related to information needs: 1) sources of information, 2) profound stress steepens the learning curve, and 3) acknowledging that learning heart failure takes time. Two categories related to experience: 4) the emotional rollercoaster, feelings of emotional distress, and 5) the hard reality of caring for a child with heart failure: always on the clock.

**Conclusions:** To our knowledge, this is the first North American situated qualitative study to provide insights about caregivers’ information needs and experiences caring for a child with chronic heart failure. This knowledge will enhance future information provision, optimizing clinical care and outcomes.

## INTRODUCTION

Chronic pediatric heart failure (PHF) is a burdensome condition for the healthcare system and families. PHF accounts for an estimated 11,000 - 14,000 hospitalizations annually in the United States.^1,2^ PHF is a factor in many chronic health conditions, such as children with cardiomyopathy, neuromuscular, metabolic, genetic, and oncologic conditions, among other chronic etiologies.^3^ A subset of children with heart failure will experience a more chronic phenotype. This population experiences systolic or diastolic dysfunction, manifesting with cardinal symptoms varying across age groups and health conditions with limited surgical treatments.^3,4^ Children with chronic heart failure and their families face an uncertain and burdensome trajectory with a tendency for repeated exacerbations requiring prolonged, specialized home and hospital care.^4^ The complexity of this situation forces families to shift their routines and learn new information quickly to provide care.

Over the past two decades, PHF has gained tremendous recognition, and as a result, treatment recommendations have evolved. Development of evidence-based treatment strategies guiding daily management has resulted from health professionals’ collaboration and knowledge-sharing.^3,4,6^ Daily management for children with chronic heart failure symptoms includes frequent medication administration, vigilant fluid management, symptom recognition, clinic appointments, and specialized diets.^3,6^ When discharged from the hospital setting, implementation, monitoring, and minor adjustments of therapies fall largely on the caregivers. These care responsibilities are also in addition to physiological pediatric growth and development needs that caregivers alone can find stressful.^7^ These combined factors provide challenges for caregivers related to treatment regimes, social and financial constraints and overall family functioning.^8^

The key to improving health outcomes in children with chronic healthcare needs is actively engaging caregivers (health consumers) with effective knowledge translation strategies.^9^ However, these strategies have lagged within the realm of pediatric heart failure. For caregivers to be proficient, evidence-based information must be relevant, understandable, and timely.^10^ However, most parents report information provision to be inadequate.^10^ It is known that caregivers who have access to understandable, evidenced-based information are more empowered to make decisions regarding their child’s health care needs more confidently. Knowledge about caregivers’ information needs in specific chronic illness contexts can assist healthcare providers in better supporting caregivers in their role as care managers.^8^ It is known that a limited number of educational tools exist for parents who have a child affected by heart failure.^11^ Understanding caregivers’ information needs and experiences through qualitative exploration has been beneficial in other chronic pediatric conditions;^12,13^ however, this knowledge has not yet been explored in chronic children’s heart failure context.

This study aimed to understand caregivers’ information needs and experiences relating to caring for a child with chronic heart failure. Knowledge generated from this study will be used to design an educational tool about pediatric heart failure for caregiver audiences.

## METHODS

### Design

A qualitative description (QD) approach was used to gain insight into this poorly understood phenomenon.^14,15^ This method is best applied when straightforward participant descriptions are desired to develop interventions related to specific populations’ needs or understanding of human behaviour.^14-17^

### Recruitment/Context

Two recruitment strategies were used to ensure a purposeful, maximally diverse sample.^18^ Purposeful sampling was conducted in a tertiary care pediatric heart failure clinic and pediatric cardiology ward. This venue was ideal for recruitment as it is the main care center that provides care to children diagnosed with chronic heart failure from five Canadian jurisdictions. To supplement recruitment from the tertiary care facility, posts about the study were shared on two research-associated social media accounts (e.g., Translating Evidence in Child Health to Enhance Outcomes (ECHO) Facebook and X accounts).

Participants were recruited if they met the study inclusion criteria (Table 1) and were willing to participate in an interview with the primary researcher (CC). We estimated that our sample size would be approximately 10-20 participants to reach data redundancy.^16^

**Table 1.** Participant Inclusion/Exclusion Criteria.

| Inclusion Criteria |
| --- |
| 1. Adult caregiver(s) (> 18 years of age) who is responsible for providing daily management & financial responsibilities for a child with chronic heart failure within the last year.<br>2. A child is defined as any person (< 18 years of age)<br>3. Child must be diagnosed with heart failure is defined as American College of Cardiology (ACC)/American Heart Association (AHA) ‘category C and D’<br>a. Stage C definition is heart disease with prior or current symptoms of heart failure.<br>Typical therapies for Stage C include, but are not limited to:<br>• Diuretics and goal-directed medical therapy<br>b. Stage D definition Patients with refractory heart failure requiring advanced intervention (i.e., biventricular pacemakers, considering left ventricular assist device but not yet implanted, transplantation)<br>4. Care must be current or within the last year to limit recall bias (e.g., if the child was transplanted or dies)<br>5. Caregivers must have completed a two-week period where the child is discharged home into the outpatient setting under their care. |
| Exclusion Criteria |
| 1. Non-English speaking participant<br>2. No access to reliable internet to complete interview |

### Ethics

Our relevant research ethics board granted ethical approval before participant recruitment (ID PRO00106559, ARISE, University of Alberta).

### Data Collection

The primary researcher (C.C.) interviewed all participants to ensure consistency. An interview guide was developed by the first author (C.C.) in consultation with senior members of the research team (J. C. and S. D. S) (Table 2). Interviews were conducted through a secure Zoom account and recorded directly onto the University of Alberta’s secure Local Area Network (LAN) portal. Recorded audio files were transcribed verbatim by a professional transcription service.

**Table 2.**
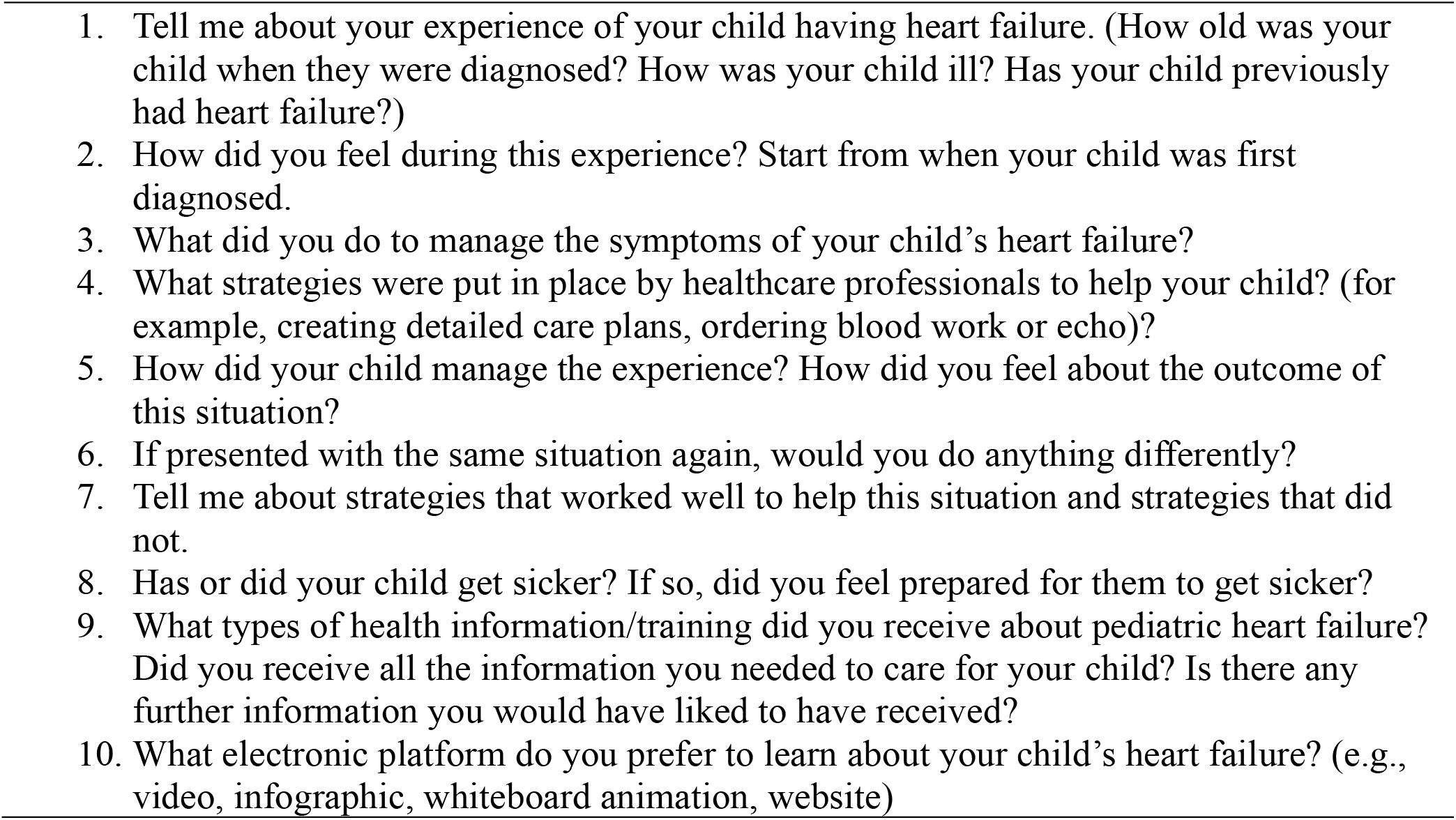
Semi-Structured Interview Guide.

Field notes were also recorded after each interview. Data collection and analysis were an iterative process to achieve data redundancy. The first author planned to contact participants up to three times via email, modelled after evidence-based surveying methods.^19^

### Analysis

Our analysis used conventional content analysis to uncover categories and sub-categories.^20^

### Rigor

Lincoln and Guba’s (1985) four trustworthiness criteria (credibility, dependability, confirmability, and transferability) guided our study.^21-23^ Credibility was established by prolonged engagement with participants during the interviews, generating thick, rich descriptions. Interview lengths ranged from 25 to 84 minutes (average 43 minutes) with large chunks of participant text.

Caregivers were highly engaged and eager to share their experiences, resulting in rich, detailed data. A third strategy to ensure rigor was peer debriefing, where a senior, experienced author-verified quotations & data warranted to establish a category. Lastly, field notes and reflexive journaling were completed following each interview to examine researcher bias. All methodological decisions were recorded. Standards for Reporting Qualitative Research (SRQR) was used to ensure comprehensive reporting.^24,25^

## RESULTS

### Demographics

Fourteen participants who identified as caregivers caring for a child with chronic heart failure participated in the study. Three participants withdrew from the study (e.g., no response via repeated email requests for an interview from the primary researcher after recruitment). Eleven caregiver interviews were conducted. Demographic data is provided in Table 3.

**Table 3.** Summary Caregiver Sample (n=11)

| Variable | Type | N |
| --- | --- | --- |
| Gender | Male | 5 |
|  | Female | 9 |
| Primary language | English | 13 |
|  | Tagalog | 1 |
| Average Number of additional adults in the home | Number of Adults | 1.9 |
| Highest level of education | College/technical school | 5 |
|  | Bachelor's degree | 5 |
|  | Post-graduate training | 1 |
| Current working status | Full-time | 9 |
|  | Part-time | 1 |
|  | Not in the Labor Force/Mat Leave | 1 |
| Annual household income | \$60, 000 - \$100, 000 | 2 |
| | \$100, 000 - \$150, 000 | 3 |
| | ≥\$150,000 | 6 |
| Health care professional experience | Yes | 6 |
|  | No | 5 |
| Average Number of children in the home (including child with heart failure) | Number of Kids | 1.9 |
| Age of the child diagnosed with heart failure | Infant (0-12 m) | 4 |
|  | Child (1-5 y) | 7 |
| Child's heart condition | Cardiomyopathy | 9 |
|  | Congenital heart disease | 2 |
| Average Number of heart surgeries | Number of Surgeries | 0.92 |
| Average Age of Heart Failure Onset | Days | 192 |
| Listed for heart transplantation<br>(Average Number of Days: 150) | Yes | 4 |
|  | No | 7 |
| Implanted ventricular assist device | Yes | 1 |
|  | No | 10 |
| Average Number of Days Since Heart Failure Diagnosis/Onset | Days | 195 |

Five categories were identified. Three categories related to caregiver information needs: 1) sources of information, 2) profound stress steepens the learning curve, and 3) acknowledging that learning heart failure takes time. The last two categories related to experience: 4) the emotional rollercoaster: feelings of emotional distress and 5) the hard reality of caring for a child with heart failure: always on the clock (Table 4).

**Table 4.** Category Labels.

| **Section #1: Information Needs** |
| --- |
| Category 1: Current Sources of Information |
| Sub-category 1. Tangible Educational Information |
| Sub-category 2. Independent Online Research |
| Sub-category 3. Healthcare Providers |
| Sub-category 4. Peer Support: Sharing Stories |
| Sub-category 5: 'Care by Parent': The Dry Run |
| Category 2: Profound Stress Steepens the Learning Curve |
| Category 3: Acknowledging that Learning Heart Failure Takes Time |
| **Section #2: Experience** |
| Category 4: The Emotional Rollercoaster: Feelings of Emotional Distress |
| Category 5: The Hard Reality of Caring for a Child with Heart Failure: Always on the Clock |

### Section #1 Information Needs

#### Category 1 Sources of Information

Caregivers described where they garnered information about their child’s heart failure. Four main sources were identified from interview transcripts: 1) tangible educational information, 2) independent online research, 3) healthcare providers, 4) peer support and sharing stories, and 5) ‘Care by Parent’: The Dry Run.

##### Tangible Educational Information

Tangible information sources are hard copies of information, like printouts or pamphlets. Participants reported that healthcare providers provided them with tangible sources, generally early on, after their child’s diagnosis. Transcript descriptions indicated that this information was usually only referred to briefly after receiving it due to several factors (e.g., misplacing their copy, only remembering a fraction of the information). Digital sources of information were more preferred as they were more convenient to access. Sorting through multiple sheets of paper or pamphlets was overwhelming and time-consuming for caregivers. Overall, this format of educational information only provided a limited amount of overall information to caregivers in the early stages. For example, one caregiver shared:

> *Um, I think it was easy to forget a lot of the stuff that was in the booklet. Once you kind of get into the thick of things, it did help to kind of understand, sort of putting some fundamentals in place. Um, and maybe like a few of those building blocks to start learning about what heart failure means*. (P1, mother)

Another caregiver shared that tangible information sources did not meet their learning needs, describing, “I didn’t find them [printouts] helpful, I actually preferred just going online and trying to do my own research online like for [child’s name] specific heart stuff.” (P11, mother).

Similarly, another caregiver shared:

> *No, generally, I find you get a ten- or fifteen-page handout at the end of the doctor visit; that’s the last thing you want to look at when I’m leaving the hospital from a visit. It’s the last thing I want to do. So, no, I don’t really enjoy those*. (P8, father)

##### Independent Online Research

Another information source described by caregivers was ‘independent’ online resources. They found they could fill specific knowledge gap(s) at specific times by tailoring their search strategies to find what they needed. This source of information was termed ‘independent’ as caregivers could complete it alone, at their own pace most convenient for them to learn, using their own search methods. Descriptions of strategies using a common search engine were often referred to by participants, *“*I just Googled stuff. That was our main source of getting that information*”* (P4, father).

Interestingly, a caregiver described that they used a common search engine and would quickly discern relevant information during their online search. They said:

… *there’s such a gap in information, like I remember, kind of in those early days when we [spouse and participant] were still trying to understand*… *I would sit in the evening, we’d be like, I Googled it, did you Google it? And they’re like, yeah, <Laughs>. I know you have to kind of like parse out the, you know, the stuff you don’t know, ‘cause we’d be like, well, that doesn’t really apply. So, we’re kind of just gonna take it with a grain of salt…*(P1, mother)

Participant observations about their partner conducting online searches were also described, highlighting the convenience of online searches, the stress caregivers endured and their desperate desire for information. One caregiver explained, “[my spouse] was literally staying up, I don’t know what hours of the night I could see [them]. I’d roll over to my bedsides and see [my spouse] looking at their phone doing research” (P4, father).

Caregivers described how their online searches were tailored to find information pertaining to current challenges or stressors they faced about their child’s heart failure. One caregiver expressed:

> *And with feeding, we had to do so much independent research, on our own about offering food…* (P3, mother)

##### Healthcare Providers

Healthcare professionals’ teaching and support were described as a valuable source of information. For example, a caregiver described their positive engagement with health professional experience, which fostered information uptake.

> *Our team has been helpful in making sure I know to ask a lot of questions too and keep a log. I want to make sure that I know what I’m looking for. Our team has really helped prepare us for something to go downhill, if it were to happen*. (P11, mother)

Caregivers with a healthcare provider background were also deemed valuable during the learning phase. One caregiver illustrated their need for extra support from the nursing staff due to this experiential knowledge gap compared to his spouse, who trained as a healthcare professional. They expressed, “It was easier for [spouse’s name] because [they are] a nurse, but I had to repeat it again, and again, and again until I was comfortable before they allowed my child to go home*”* (P10, father).

##### Peer Support: Sharing Stories

Important connections and sharing of stories helped caregivers better understand their child’s heart failure while also helping them learn and discover new, strategic ways of navigating the healthcare system. Relating to other caregivers in a similar context was described as helpful to caregivers and fostered coping. One caregiver shared:

> *You know, so you hear anecdotally from other families who are like, “Oh no, no, no, no*.*” We were told that this would never happen, but then it did. And now, you know, here’s our life now. We now feed our kid with a pump…. So yeah, I think maybe, especially in the beginning, when you’re still kind of in that overwhelming phase of, um, just drinking from the fire hose in terms of information, a peer support group of someone kind is helpful*. (P1, mother)

Similarly, two caregiver descriptions also valued having peer support:

> *Um, I remember, uh, [the physician] put me in contact with another family whose little girl has cardiomyopathy with heart failure, who is currently on the transplant list. It was so helpful to sit down and talk to [caregiver name]. Their experience was different than mine and has turned out different than mine, but*… *you’re talking to somebody who understands, who understands hospital life, they understand trying to navigate this, this condition that you’ve never previously known about…* (P1, mother)
>
> *…someone that you could relate to, who are going through the same troubles. You know, and, and they would talk about their child, and you’ll talk about yours. And it was just a good common ground to relate to somebody else, you know* (P4, father)

Caregivers valued hearing stories of survival. This provided them with hope from families whose children survived. For example, one caregiver stated, “I was reading up on others’ experiences with heart transplant. I found that to be really helpful” (P11, mother). Another caregiver described, “…I connected with some other families dealing with the same situation. That kind of helped a little bit. Some kids were surviving” (P10, father).

##### “Care by Parent’: The Dry Run

‘Care by Parent’ is a dry run of daily care tasks completed by responsible caregivers in the hospital environment at the end of a pediatric patient’s stay. Caregivers described how this pragmatic learning method in the hospital improved their confidence. One caregiver shared how they liked the “hands-on approach or giving of examples by healthcare staff” (P4, father). Another caregiver shared how it gave them the “opportunity to ask questions.” (P6, mother). Other participants described:

> *I would say that it [care by parent] was really valuable to us. I think a lot of the care that we learned how to do before we came home, we learned in hospital. We learned that through the nurses giving the care to [our child], so we were able to really easily pick that up when we were at home*… (P3, mother)
>
> *… So just like measuring her meds, they came in three or four times to make sure we were doing it right. They let us do it a few times to the point where we were comfortable. Um, gave us everything to look for in case she needed to come back, like she’s retaining water is a big one, her color, all that information…*. (P5, mother)

A caregiver described how doing a dry run increased their overall confidence. They shared, “They taught us the essential basics, but at the same time, it made us feel confident. We were ready to go home is what we felt” (P7, father).

#### Category #2 Profound Stress Steepens the Learning Curve

Hearing that their child was diagnosed with heart failure was overwhelmingly debilitating for caregivers. This affected their ability to effectively uptake and understand information following the news of a new stressor. Caregivers typically experienced this inability to understand new information after initial diagnosis or if their child had a decline in health, both being at times of heightened stress. One caregiver illustrated this by sharing:

> *Um, so I think that the learning curve has been steep. On top of dealing with, accepting, and coming to terms with the fact that [child’s name] has heart failure, you’re also simultaneously moving along that learning curve, which you know, you’re being thrown in the pool, in the deep end of the pool, right?* (P1, mother)

Two other caregivers described this phenomenon:

> *Because when you get a new diagnosis, you can’t think. It’s like time freezes. You’re observing your life instead of living it. And it’s hard, it’s hard to remember <Tearing up>… But, in the beginning, I felt like we were probably just a deer, deer in the headlights, like totally overwhelmed being first-time parents and then even more overwhelmed by the fact we have a kid that was really sick and, uh, trying to navigate that… And so, the rest was kind of a blur after that. No one really explained to us what [heart failure] was really. They kind of explained to us, but it didn’t really make any sense, like why we were there? What was happening? We didn’t know. And they asked us all these questions and none of it made sense*. (P4, father)
>
> *If there is a new diagnosis, somebody came in and talked about all the facets of what that means. Sometimes at rounds, people are just like spitting things off and you don’t really, you don’t really understand, like, I heard that they were gonna give her formula and it made me really emotional. Like, I was very upset, but now obviously looking back [our child] had chylothorax and needed the formula* (P3, mother)

#### Category 3 Acknowledging that Learning Heart Failure Takes Time

Caregivers discussed that understanding information about their child’s heart failure eventually became easier over time. Factors that allowed caregivers to better uptake information over time were lessened stress levels and repeated exposure to information. One caregiver describes this experience:

> *Now, I feel like every time she has a blip or gets sick, I start to realize – I can see it better after, once she’s doing better and I reflect back, then I see more clearly kind of what was happening or that it was, you know, if she was sick because of an infection or just sick in general like a normal kid. Looking back, you can see how hard it probably was – or it was on her heart failure as well, whereas during the time, you don’t see it*. (P9, mother)

Similarly, other caregivers described that repeated exposure to information was helpful:

> *…I don’t think [feeding] quite clicked until there was an online feeding therapy conference. And I watched like the first 30 minutes of that and just like cried <Laughs> and I was like, “Oh my god, I probably made my kid sicker, like…*.*” It was really hard to wrap my head around that before, because you know, you think about medicine. And then you think about what role does diet play. I think just having a better understanding of the severity was really… I just wish I would’ve understood the repercussions of that when it was happening because I think um, we probably would’ve done things a little bit differently if that was the case*. (P3, mother)
>
> *So, I think over time it [learning about their child’s heart failure] got better. I got a better understanding and that learning curve was a little less steep*… *<Laughs>… I had like no concept of what was being talked about, and I didn’t even know where to start asking questions. Um, so I would say the biggest challenge would be that I didn’t know what I didn’t know. And so when I was asked, you know, well, “What questions do you have?” And it’s like, “I probably have a thousand questions but I just don’t know what they are*. (P1, mother)

Another caregiver also shared a similar experience:

> *Yeah, just too like re-go back to it, to reference it, if that make sense. Especially as you go through learning, so when you’re given some information in the hospital, it doesn’t make a whole lot of sense at first, but then as you, as time progresses, when you revert back to it, it’s like, oh yeah, this makes sense*… (P4, father)

One caregiver described how repeated exposure to information over time allowed them to retain smaller components of complex information slowly. They shared, “Oh, that’s what they’ve been talking about this whole time. Okay. This all makes, okay. This little piece makes sense. Now I’m gonna store that away. And now I know something a little bit more” (P1, mother).

### Analysis Section 2: Experience

#### Category 4 The Emotional Rollercoaster: Feelings of Emotional Distress

All caregivers described accounts of experiencing emotional distress. The distress was highest following diagnosis due to the severity and shock of such a severe diagnosis. For example, caregiver descriptions included the feelings of “denial” (P3, mother), “loss of control” (P4, father), or the situation being “overwhelming” (P1, mother), or “difficult to accept” (P10, father). One caregiver explained that their “whole world was turned upside down” (P6, mother). Caregivers also shared that it was difficult to hear such traumatic information about their child. One example is a participant stating, “I don’t think you can ever be completely prepared” (P11, mother).

Several caregivers also described feeling emotionally distressed from the uncertainty of their child’s survival. One caregiver stated, “Uncertainty is the hardest part” (P9, mother). Another caregiver described:

> *He’s very sick. He’s dying really to put it bluntly. And I know that a transplant too is just a Band-Aid. He’s always going to be sick. And then there’s a risk later on too that he would reject that heart and he needs another transplant. There’s definitely days where it’s a lot harder and it kind of hits all at once*. (P11, mother)

Another caregiver shared their feelings of uncertainty as they expressed, “The other part that wasn’t easy was we never knew if what was going to be the outcome. We didn’t know if [our child] was going to survive the situation or not.” (P10, father)

Caregivers also described feelings of anger and denial when their child’s health worsened. One participant shared:

> *And fast forward here a year, um, when [Daughter]’s heart function plummeted, I found it very hard to understand what was happening. Because so many people were saying, um, she needed a heart transplant. And I remember, I think it was [Doctor] came in and really hounded us that she needed a heart transplant. And I got mad at [doctor], I got really mad*… (P4, father)
>
> Another example of participants feeling emotional distressed related to descriptions of denial. One caregiver stated:
>
> *I think [my spouse] has definitely been more accepting and understanding of the situation. Whereas I have been like, “She’s not sick, she doesn’t need this*.*” Like even, um, you know, when we did the write-up for transplantation. Like, I couldn’t wrap my head around the fact that she needed a heart transplant, and I still, to some degree, don’t really believe it <Shaking Head>*. (P2, father)

#### Category 5 The Hard Reality of Caring for a Child with Heart Failure: Always on the Clock

Several caregivers described difficulties managing their child’s complex heart failure treatment schedule. This resulted in stress and constant self-sacrifice from always having to be on the clock. Caregivers shared negative feelings about constantly having to complete regimented daily tasks, highlighting how it impacted their health and family functioning. For example, one caregiver described:

> *Physically, [my child] is doing fantastic. They are on five different types of medications, not including vitamin D drops. Um, that’s been slightly difficult. I feel like I’m torturing [my child], even though I know I’m really not <Laughs>*. (P5, mother)

Caregivers described challenges relating to a cumbersome, regimented schedule, describing their daily parenting routine as ‘following an instruction manual.’ They shared:

> *Because she was given such strict feeding volumes, feeding schedule and meds schedule and everything. So it was it, you know, off the start, it really felt like we were, you know, I felt like I’m just the whole time kind of following an instruction manual right? Of, you know, things have to step one, step two, step three all throughout the day and then repeat, right? And it, um, you know, made it very hard to, um, kind of do anything normal, right? … So even, even just wanting to, you know, get out of, get out of there, um, for a day, or so go to the mall, made it kind of difficult. It was a lot of planning because we had to make sure we had her meds and we had her food and we had a way warm in and you know, all that. So, um, it just, just very regimented I guess, was, was the experience off the start*. (P2, father)

Another caregiver shared how there was also feelings of anxiety related to meeting their child’s fluid requirements. They stated, “Um, well, actually, when she was very young, she wasn’t too bad cause she would finish most of her bottles. Um, but it was always sort of this anxiety around, um, you know, if she doesn’t.” (P1, mother)

Another caregiver described similar feelings but expressed anger when the prescribed feeding volumes were unmet.

> *So, I just found myself getting, you know, frustrated and even angry sometimes when she would have a bad day. And it’s like, whereas with our, you know, say with our son, it’s like, whatever, he didn’t eat today, he just wasn’t hungry. Who cares? Right? He’ll do good the next day, but that was never the case with her. …And I know she’s not doing as good as she could that it almost makes me, I’ll say, get angry at her, but I don’t know if that’s really right. I mean, she’s the focus of things. And if I’m angry, she’s the reason. <Laughs>* (P2, father)

Caregivers also described instances where the regimented home regime strained the caregiver-child relationship. The description below highlights how the caregiver compares water to heroin due to the constant manipulation from their child, signifying the strain caregivers feel when trying to balance prescribed care regimes. They shared:

> *I think she knows she can’t manipulate me as easily, so I don’t think I’m her biggest target in that, in that sense. I think if she wants water <Laughs> she will ask her dad. To be honest with you. Uh, which also is like really funny, how she’s already figured out that like, dad will bend, mom will not. So let’s go, where we know our, our effort will be recognized, I guess… It’s like having a little heroin addict but like the water is the heroin. Cause she’s just like “water, water?” And she will manipulate the hell out of you to get water*. (P3)

## DISCUSSION

Our findings build on very limited published knowledge about caregivers’ information needs and experiences caring for a child with chronic heart failure. Our paper provides new findings from a North American lens and provides a more in-depth understanding of caregiver information needs. Zhang et al. (2023) published a study from Asia that uncovered findings related to challenges with family management with a child with chronic heart failure, uncovering three themes.^26^ The themes were titled 1) weakened family socialization, 2) the experience of five psychological stages, and 3) family management dilemmas. Each theme had 2-5 subthemes supporting the development of the main theme. Elements of these themes can certainly be relatable in our 5 themes, namely our emotional distress category and Zhang (2023) psychological stages theme (worry and exhaustion).^26^ Our paper provides new knowledge concerning parent learning, whereas Zhang suggests insufficient information was available under their family management dilemma theme but does not provide any further detail.

Since there are no other pediatric chronic heart failure studies available for comparison, a rapid review in 2015 synthesized 34 included studies of parents who had a child with a chronic illness.^8^ Comparing the chronicity of PHF to other health conditions would likely have some similarities relating to parents’ information needs and experiences. Of the chronic illnesses, no caregivers who cared for a child with heart failure were included in this study. Three main themes were identified in Smith’s paper: (1) caregiver impact: making sense of the condition, grief and loss; (2) illness management: learning about the condition, monitoring symptoms, and responding to changes in the child’s condition, interacting with health professionals; and (3) social disruption: managing disruption. Smith’s themes resonated with our five information needs and experience-themed categories but in a more generated context of pediatric chronic illness.

### Caregiver Learning and Information Needs

This study demonstrates caregiver challenges related to information uptake and retention after exposure to a stressful situation. Compelling evidence relating to congruent learning and testing environments by Schwabe and Wolfe (2009) suggests key points related to our study.^27^ They state that integrating and storing new information into an individual’s memory happens during learning. Learning can be impaired if an individual is exposed to a stressful event before undergoing the learning activity,^27^ as caregivers described in our study. We noted in our first category related to information needs, that caregivers are exposed to an extreme stressor (e.g., being told their child has heart failure). This stressor immediately impairs the caregivers’ ability to retain or place new information into their memory following that stressful event. Hence, there is a need for repetition through digital platforms, such as online formats, to help them eventually learn and retain information related to their child’s heart failure.

Secondly, Schwabe and Wolf (2009) also highlight context-dependent memory,^27^ as seen in this study. They suggest that individuals who demonstrate memorization of information in one environment (e.g., hospital) may be unable to recall and apply it in another environment (e.g., home), also known as testing. In our study, we noted that caregivers described feelings of confidence while receiving care from caregivers in the hospital environment before discharge. Caregivers had a positive experience recalling the knowledge they learned about caring for their child. However, when they returned to their home environment, the caregivers described experiencing heightened stress.

Caregivers also preferred digital educational tools as a source of learning compared to tangible educational materials. These tools provide a portable source of relevant information that could be repeated as needed. These tools also provide easy access to certain types of information (e.g., floating menu tabs with headings for topics) to access specific types of information conveniently. Most caregivers have access to a mobile device, compared to tangible educational resources (e.g., paper copies), which caregivers found cumbersome to sort through and did not provide a constant source of information (e.g., misplaced after some time). Knowledge translation strategies that employ a digital arts-based approach alleviate these issues for caregivers, fostering more effective health consumers better equipped to make decisions.^28^

### Clinical Implications

Two key clinical implications exist. Knowledge gained from this research provides clinicians with insight into families’ experiences and how to improve the provision of information to this audience (knowledge translation). Providing information that can be repeatedly accessed can help caregivers learn complex knowledge under stressful circumstances. Furthermore, as caregivers learn in different ways and at different paces, having interactive and accessible knowledge will help facilitate learning in a diverse group. Lastly, given our findings, we must recognize that caregivers will have a difficult time learning about their child’s heart failure in the early stages and supporting them during this time of steep learning with relevant and understandable education tools, healthcare provider support and practice in the clinical setting is of utmost importance to improving outcomes in this population.

### Next Steps/Future Research

The next step is to design and refine an educational tool about pediatric heart failure with caregivers. The design and content of the educational tool will be based on a combination of knowledge from the findings of caregivers who have lived experience caring for a child with heart failure and evidence-based treatment guidelines. This tool is unique in that the design is based upon qualitative interviews about caregivers’ information needs experience in its design since evidence has been so limited.

Future research could also expand on the work done here by interviewing a larger cohort of caregivers throughout North America about their information needs and experiences. This would add to the knowledge base of this important topic, helping improve knowledge translation to caregivers.

### Limitations

One limitation is that all the participants were recruited from a single center, perhaps affecting the overall external validity.

The primary researcher in this study was also one of the clinicians on the family’s care team. Several steps were identified beforehand to ensure that ethics and rigour were withheld^29,30^ (e.g., the primary researcher did not approach families about participation, the research gift card was offered after the participants agreed to participate, interviews were completed in a virtual setting away from the clinical environment, review about confidentially took place before the interview occurred to ensure the participants were reminded about the ethical principles designed to protect participants).

## CONCLUSION

Our qualitative interviews have uncovered 5 categories. Three categories relate to caregiver information needs: 1) sources of information, 2) profound stress steepens the learning curve, and 3) acknowledging that learning heart failure takes time. The last two categories relate to caregiver experience: 4) the emotional rollercoaster: feelings of emotional distress and 5) the hard reality of caring for a child with heart failure: always on the clock. Similar to a previously published study, our study adds to a limited foundation of knowledge about parental emotional distress and lack of available information but adds findings about challenges with information uptake and retention and lack of information that caregivers face in this difficult situation. Our study was a much-needed step in improving the provision of information to caregivers through research knowledge. Furthermore, our paper will apply this knowledge in the design of a digital education tool for caregivers.

## Data Availability

The data that support the findings of this study are available from the corresponding author upon reasonable request.

## Acknowledgements

The authors of this study would like to thank all the participants who shared their impactful experiences with the research team.

CC is funded by the Canadian Institute of Health Research Doctoral Fellowship (RES0056719), a previous graduate studentship from the generous support of the Women’s and Children’s Health Research Institute (WCHRI), numerous awards from the Alberta Registered Nurse Educational Trust (ARNET), the University of Alberta Faculties of Nursing and Graduate Student and Postdoctoral Studies (GPS). ZZ was generously funded by the Women’s and Children’s Health Research Institute (WCHRI). SS is a Distinguished Researcher at the Stollery Children’s Hospital (RES0044689). SS is also funded by CIHR (RES RES0044689).

## Ethical Approval

This study was approved by the University of Alberta Ethics Board (ARISE ID Pro00106559).

## Statement of Human and Animal Rights

All procedures in this study were conducted in accordance with the approved protocols of the University of Alberta Ethics Review Board (ARISE ID Pro00106559).

## Statement of Informed Consent

Written informed consent was obtained from the patient(s) for their anonymized information to be published in this article.

## Notes

### Competing Interest Statement

The authors have declared no competing interest.

### Clinical Trial

N/A

### Funding Statement

Canadian Institute of Health Research (CIHR) Women and Children's Health Research Institute (WCHRI) Alberta Registered Nurses Educational Trust (ARNET)

### Author Declarations

University of Alberta Health Ethics Board (ARISE, ID PRO00106559)

## REFERENCES

1. Amdani S, Marino B, Rossano J, Lopez R, Schold J, Tang, W. Burden of pediatric heart failure in the United States. J Am Coll Cardiol. 2022;79(19):1917–1928. doi: 10.1016/j.jacc.2022.03.336

2. Rossano JW, Kim JJ, Decker JA, Price JF, Zafar F, Graves DE, Morales DL, Heinle JS, Bozkurt B, Towbin JA, … Jefferies, J. L. Prevalence, morbidity, and mortality of heart failure-related hospitalizations in children in the United States: A population-based study. J Card Fail. 2012;18(6): 459–470. doi: 10.1016/j.cardfail.2012.03.001

3. Kantor PF, Lougheed J, Dancea A, McGillion M, Barbosa N, Chan C, Dillenburg R, Atallah J, Buchholz H, Chant-Gambacort C, … Children’s Heart Failure Study, Presentation, diagnosis, and medical management of heart failure in children: Canadian Cardiovascular Society guidelines. Can J Cardiol. 2013;29(12): 1535–1552. doi: 10.1016/j.cjca.2013.08.008

4. Das BB. Current state of pediatric heart failure. Child. 2018;5(7):88 doi: 10.3390/children5070088

5. Bansal N, Burstein DS, Lorts A, Smyth L, Rosenthal DN, Peng DM. Heart failure in children: priorities and approach of the ACTION collaborative. Prog Pediatr Cardiol. 2020;59:101313. doi: 10.1016/j.ppedcard.2020.101313

6. Kirk R, Dipchand AI, Mertens L, Rosenthal DN, Dubin A, Addonizio L, … Weintraub R. The International Society for Heart and Lung Transplantation guidelines for the management of pediatric heart failure: executive summary. [Corrected]. J Heart Lung Transplant. 2014;33(9):888–909. doi: 10.1016/j.healun.2014.06.002

7. Kepreotes E, Keatinge D, Stone T. The experience of parenting children with chronic health conditions: a new reality. J Nurs Healthcare Chronic Illnesses. 2010;2(1):51–62. doi: 10.1111/j.1752-9824.2010.01047.x

8. Smith J, Cheater F, Bekker H. Parents' experiences of living with a child with a long-term condition: a rapid structured review of the literature. Health Expect, 2015;18(4), 452–474. doi: 10.1111/hex.12040

9. Hartling, L, Elliott SA, Buckreus K, Leung J, Scott SD. Development and evaluation of a parent advisory group to inform a research program for knowledge translation in child health. Res Involvement Engage. 2021;7(1):38. doi: 10.1186/s40900-021-00280-3

10. Hummelinck A, Pollock K. Parents’ information needs about the treatment of their chronically ill child: A qualitative study. Patient Education Counsel. 2006;62(2):228-234-234. doi: 10.1016/j.pec.2005.07.006

11. Cunningham C, Sung H, Benoit J, Conway J, Scott SD. Multimedia knowledge translation tools for parents about childhood heart failure: environmental scan. [Corrected]. JMIR Pediatr Parent. 2022;5(1):e34166. doi: 10.2196.34166

12. Gates M, Shulhan-Kilroy J, Featherstone R, MacGregor T, Scott SD, Hartling, L. Parent experiences and information needs related to bronchiolitis: a mixed studies systematic review. Patient Education Counsel. 2019;102(5), 864–878. doi: 10.1016/j.pec.2018.12.013

13. Meherali S, Campbell A, Hartling L, – Scott S. Understanding parents’ experiences and information needs on pediatric acute otitis media: a qualitative study. J Patient Experience. 2019;6(1):53–61. doi: 10.1177/2374373518771362

14. Sandelowski M. Focus on research methods: whatever happened to qualitative description? Res Nurs Health. 2000;23(4):334–340. doi: 10.1002/1098-240x(200008)23:4<334::aid-nur9>3.0.co;2-g

15. Sandelowski, M. What’s in a name? Qualitative description revisited. Res Nurs Health. 2010;33(1):77–84. doi: 10.1002/nur.20362

16. Kim H, Sefcik JS, Bradway C. Characteristics of qualitative descriptive studies: a systematic review. Res Nurs Health. 2017;40(1):23–42. doi: 10.1002/nur.21768

17. Neergaard M, Olesen F, Andersen R, Sondergaard J. Qualitative description – the poor cousin of health research? BMC Med Res Methodol. 2019;9(1):52. doi: 10.1186/1471-2288-9-52

18. Lee-Jen Wu S, Hui-Man H, Hao-Hsien L. [A Comparison of Convenience Sampling and Purposive Sampling]. J Nurs. 2014;61(3):105–111. doi: 10.6224/JN.61.3.105

19. Dillman DA. (2007). Mail and internet surveys: The tailored design method, 2nd ed. John Wiley – Sons Inc.

20. Hsiu-Fang H, Shannon SE. Three approaches to qualitative content analysis. Qual Health Res. 2005;15(9):1277–1288. doi: 10.1177/1049732305276687

21. Lincoln YS, Guba EG. (1985). Naturalistic inquiry. Sage Publications.

22. Morse JM. Critical analysis of strategies for determining rigor in qualitative inquiry. Qual Health Res. 2015;25(9):1212–1222. doi: 10.1177/1049732315588501

23. Stahl NA, King JR. (2020). Expanding approaches for research: understanding and using trustworthiness in qualitative research. J Devel Education. 2020;44(1): 26–28. http://www.jstor.org/stable/45381095

24. O’Brien BC, Harris IB, Beckman TJ, Reed DA, Cook DA. (2014). Standards for reporting qualitative research: a synthesis of recommendations. Acad Med. 2014;89(9):1245–1251. doi: 10.1097/ACM.0000000000000388

25. Dossett LA, Kaji, A., Cochran A. SRQR and COREQ reporting guidelines for qualtiative studies. JAMA Surg. 2021;156(9):875–876. doi: 10.1001/jamasurg.2021.0525

26. Zhang A, Zheng Z, Shen Q, Zhang Q, Leng H. Family management experience of children with chronic heart failure: a qualitative study. J Pediatr Nurs. 2023;73:e36–42. doi: 10.1016/j.pedn.2023.07.006

27. Schwabe L, Wolf OT. (2009). The context counts: congruent learning and testing environments prevent memory retrieval impairment following stress. Cogn, Affective, Behav Neurosci. 2009;9(3):229–236. doi: 10.3758/CABN.9.3.229

28. Albrecht L, Scott SD, Hartling L. (2017). Knowledge translation tools for parents on child health topics: a scoping review. BMC Health Serv Res. 2017;29:686. doi: 10.1186/s12913-017-2632-2.

29. Yanos PT, Ziedonis DM. The patient-oriented clinician-researcher: advantages and challenges of being a double agent. Psychiatr Serv. 2006;57(2):249–253. doi: 10.1176/appi.ps.57.2.249

30. Largent EA, Lynch HF. Paying research participants: regulatory uncertainty, conceptual confusion, and a path forward. Yale J Health Policy, Law – Ethics. 2017;17(1):61–141.

